# HNK-1 Antibody-Based ELISA for Soluble PTPRZ: A Practical Cerebrospinal Fluid Biomarker for Glioma

**DOI:** 10.1101/2025.08.19.25332842

**Authors:** Yu Naruse, Masazumi Fujii, Kenichiro Nagai, Ryo Hiruta, Toshie Sakagami, Takuru Kobayashi, Kazuto Takahashi, Junko Iijima, Hideaki Suzuki, Kasumi Hattori, Kazuaki Kanai, Yuka Oka, Yuko Hashimoto, Shunichi Koriyama, Taiichi Saito, Yoshihiro Muragaki, Takakazu Kawamata, Takashi Sasayama, Jun Yasuda, Miwa Uzuki, Yasushi Kawaguchi, Kentaro Kawata, Yoshiki Yamaguchi, Shiori Go, Hiromu Arakawa, Hiroyuki Kaji, Shinobu Kitazume

## Abstract

Reliable fluid biomarkers for glioma remain elusive, complicating diagnosis and differentiation from primary central nervous system lymphoma (PCNSL). We evaluated soluble protein tyrosine phosphatase receptor type Z (sPTPRZ) in cerebrospinal fluid (CSF) using an anti-HNK-1 antibody-based ELISA. CSF sPTPRZ levels were significantly elevated in glioma patients compared with controls (AUC = 0.910) and moderately distinguished gliomas from PCNSL (AUC = 0.805). Immunohistochemistry and qPCR confirmed lack of PTPRZ expression in PCNSL, while epitope mapping localized the HNK-1 modification site to exon 12 of the long isoform. RNA-seq of the C-CAT database revealed PTPRZ-MET fusions in 2% of gliomas, all lacking the HNK-1 epitope, consistent with low CSF sPTPRZ in a subset of tumors. These findings establish CSF sPTPRZ detection with anti-HNK-1 antibody as a minimally invasive and robust biomarker for glioma, supporting improved diagnostic accuracy and aiding clinical decision-making in neuro-oncology.

## Introduction

Gliomas are among the most common primary brain tumors and remain a challenging group of intractable cancers with significant obstacles in both diagnosis and treatment^1–3^. Among the various challenges in clinical management, the absence of specific and clinically applicable fluid-based diagnostic biomarkers is, especially, a critical issue. Because imaging studies alone often fail to provide a definitive diagnosis, invasive procedures such as biopsy are sometimes required. To enable timely and accurate diagnosis that facilitates prompt therapeutic intervention, the development of non-invasive and rapid fluid-based diagnostic methods is urgently needed. Moreover, current imaging modalities are often insufficient for reliably assessing treatment response or detecting recurrence. Therefore, the identification of specific fluid biomarkers is highly desirable for improving disease monitoring and clinical decision-making.

PTPRZ is a membrane-bound protein that is highly expressed in central nervous system (CNS) glial cells, including oligodendrocyte precursor cells, astrocytes, and oligodendrocytes^4,5^ . It has been shown that PTPRZ is more highly expressed in glioma cells compared to normal brain tissue ^6^ ^7^ ^8^. Furthermore, the extracellular region is cleaved and shed, and the soluble form is known as sPTPRZ is markedly elevated in the cerebrospinal fluid (CSF) of glioma patients^9^, suggesting that sPTPRZ may serve as a promising biomarker candidate for gliomas.

PTPRZ is heavily glycosylated with chondroitin sulfate ^10^, keratan sulfate ^11^, N-glycans, and unique human natural killer-1 (HNK-1) epitope consists of a sulfated trisaccharide ^12^ is attached to the O-mannosyl core M2 glycans ^13,14^. So far, HNK-1-capped O-mannosyl glycans and core M2 glycans are only found in PTPRZ^15^. In mice deficient in GnT-IX, a glycosyltransferase involved in the formation of the core M2 branching structure, a reduction in the HNK-1 epitope on PTPRZ has been observed^14^. Furthermore, in a xenograft model using glioma cells with

GnT-IX knockdown, a marked reduction in tumor growth was observed, indicating that the core M2 glycan carrying the HNK-1 epitope on PTPRZ plays a regulatory role in glioma tumorigenicity^16^. PTPRZ has been implicated in glioma progression and may serve as a marker or therapeutic target for glioma ^9,16,17^. However, the detailed expression levels of sPTPRZ in the cerebrospinal fluid (CSF) remain unclear for other types of brain tumors, including primary central nervous system lymphoma (PCNSL), which is of particular clinical importance in the differential diagnosis of gliomas.

In this study, we evaluated whether CSF sPTPRZ could be useful in distinguishing glioma from PCNSL. To do this, we have developed sandwich ELISA with anti-HNK-1 antibody, which is shown to be specifically and highly reactive with CSF sPTPRZ. Our findings revealed that CSF sPTPRZ derived from tumors was more abundant in glioma patients than in controls and PCNSL patients. This study suggests that CSF sPTPRZ has potential as a liquid biopsy marker for minimally invasive differentiation between glioma and PCNSL.

## Results

### Analysis of CSF sPTPRZ with anti-HNK-1 antibody

MRI findings for gliomas and PCNSLs can sometimes overlap, making differential diagnosis challenging. Figure 1A shows MRI images of a typical PCNSL and a glioblastoma with MRI findings resembling those of PCNSL. The MRI images of glioblastoma demonstrated hypo-intense on T1-weighted imaging (T1WI), hyper-intense on T2-weighted imaging (T2WI), hyper-intense on diffusion-weighted imaging (DWI), hyper-intense on fluid attenuated inversion recovery (FLAIR) imaging, and homogenous enhancement on gadolinium enhanced imaging, requiring a tissue biopsy for differentiation. To clarify if CSF sPTPRZ could be useful in distinguishing glioma from PCNSL, we first performed western blotting using CSF from glioma and PCNSL patients, and control samples. Most of commercially available anti-PTPRZ antibodies do not detect heavily glycosylated CSF sPTPRZ and limited anti-PTPRZ antibodies, such as those from Santa Cruz, require chondroitinase ABC (ChABC) treatment to enable detection, and these anti-PTPRZ IgM antibodies recognize the HNK-1 epitope on the O-mannosyl glycans of sPTPRZ^16,18^. Therefore, we here first examined the utility of the anti-HNK-1 IgG antibody (anti-SGPG, TCI Chemicals)^19^ for detecting CSF PTPRZ. When CSF from the same glioma patient was tested, the anti-HNK-1 antibody displayed a stronger signal as compared with the anti-PTPRZ (Santa Cruz) antibody (Fig. 1B). The anti-HNK-1 antibody showed a signal even in CSF without ChABC treatment. We also confirm that the anti-HNK-1 antibody dose recognize sPTPRZ, since the CSF immunoprecipitants with the anti-HNK-1 antibody were clearly detected with the anti-PTPRZ (Santa Cruz) antibody, in which the signal intensity of the immunoprecipitants was equivalent to 100% of the input (Fig. 1C). In the brain, α-amino-3-hydroxy-5-methyl-4-isoxazolepropionic acid (AMPA)-type glutamate receptor (AMPAR) subunit GluA2 is another membrane-bound glycoproteins bearing the HNK-1 epitope on the N-glycans^20,21^. However, since no distinct signal was observed around 100 kDa, corresponding to GluA2, it is considered that shedding of HNK-1-bound GluA2 into the cerebrospinal fluid (CSF) is negligible. These findings indicate that the anti-HNK-1 antibody has high affinity to CSF sPTPRZ, and we next used anti-HNK-1 and anti-PTPRZ (Santa Cruz) antibodies to evaluate sPTPRZ levels in CSF samples from glioma, PCNSL, and control groups. Strong sPTPRZ signals were observed in 3 out of 4 glioma samples with both antibodies, whereas in the control and PCNSL samples sPTPRZ signals were weak with anti-PTPRZ (Santa Cruz) and these signals were not even detectable with anti-HNK-1 antibody (Fig. 1D).

**Figure 1.**
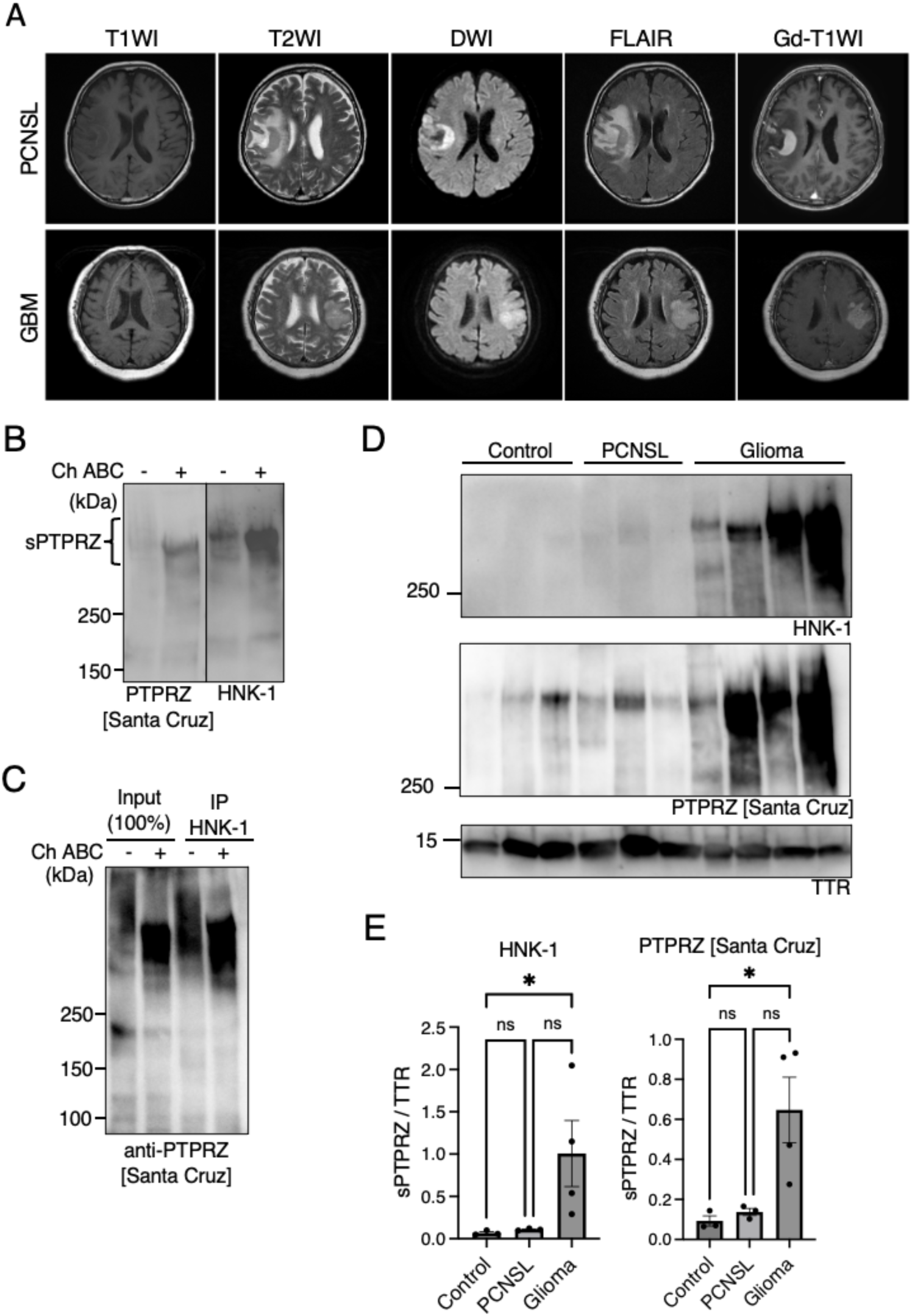
**Detection of sPTPRZ in CSF using the anti-HNK-1 antibody.** (A) Representative MRI images of a PCNSL and a glioblastoma, IDH-wildtype, WHO grade 4, with overlapping imaging characteristics. (B) Western blot analysis comparing the detection of sPTPRZ in CSF using anti-HNK-1 and anti-PTPRZ [Santa Cruz] antibodies. (C) Immunoprecipitation of sPTPRZ from CSF using anti-HNK-1 antibody, followed by detection with anti-PTPRZ [Santa Cruz] antibody. (D) Western blot analysis of sPTPRZ levels in CSF samples from glioma, PCNSL, and control patients. (E) Quantification of sPTPRZ levels in glioma, PCNSL, and control samples. Data are presented as mean ± SEM. Statistical significance was determined using the Kruskal-Wallis test.

Semiquantitative western blot analysis demonstrated that sPTPRZ levels were significantly higher in CSF from glioma patients compared with control CSF (Fig. 1E). Although sPTPRZ levels were also higher in glioma patients than in PCNSL patients, this difference was not statistically significant.

### PTPRZ protein expression in glioma and PCNSL

Low sPTPRZ levels in CSF from PCNSL patients suggests reduced PTPRZ expression in PCNSL tumor cells. To clarify this, we conducted histological examination and immunohistochemical staining of pathological sections from glioma (oligodendroglioma, IDH-mutant and 1p/19q-codeletion, WHO Grade 2; and glioblastoma, IDH-wild type) and PCNSL (diffuse large B cell lymphoma). We here used anti-HNK-1 and Cat-315 antibodies, the latter of which recognizes the HNK-1 epitope plus the PTPRZ peptide region ^18,22^. H&E staining revealed characteristic features of each tumor: glioblastomas exhibited frequent mitoses and necrosis, densely proliferating atypical cells^23^ (Fig. 2); oligodendrogliomas displayed “fried-egg”-like tumor cells^24^; PCNSLs showed diffuse infiltration of large tumor cells with round, indented, or cleaved nuclei, prominent nuclei, and CD20/CD10 positivity, CD3 negativity, and a Ki-67 index of 80%, consistent with diffuse large B-cell lymphoma^25^. Glioblastoma and oligodendroglioma tumor cells were positive for both Cat-315 and anti-HNK-1 antibodies in the cytoplasm, whereas PCNSL tumor cells showed no reaction with either antibody. Several Cat-315 and anti-HNK-1 positive signals observed in PCNSL samples were attributed to surrounding glial tissue. These findings indicate that gliomas express PTPRZ with the HNK-1 epitope, which is absent in PCNSL tumor cells.

**Figure 2.**
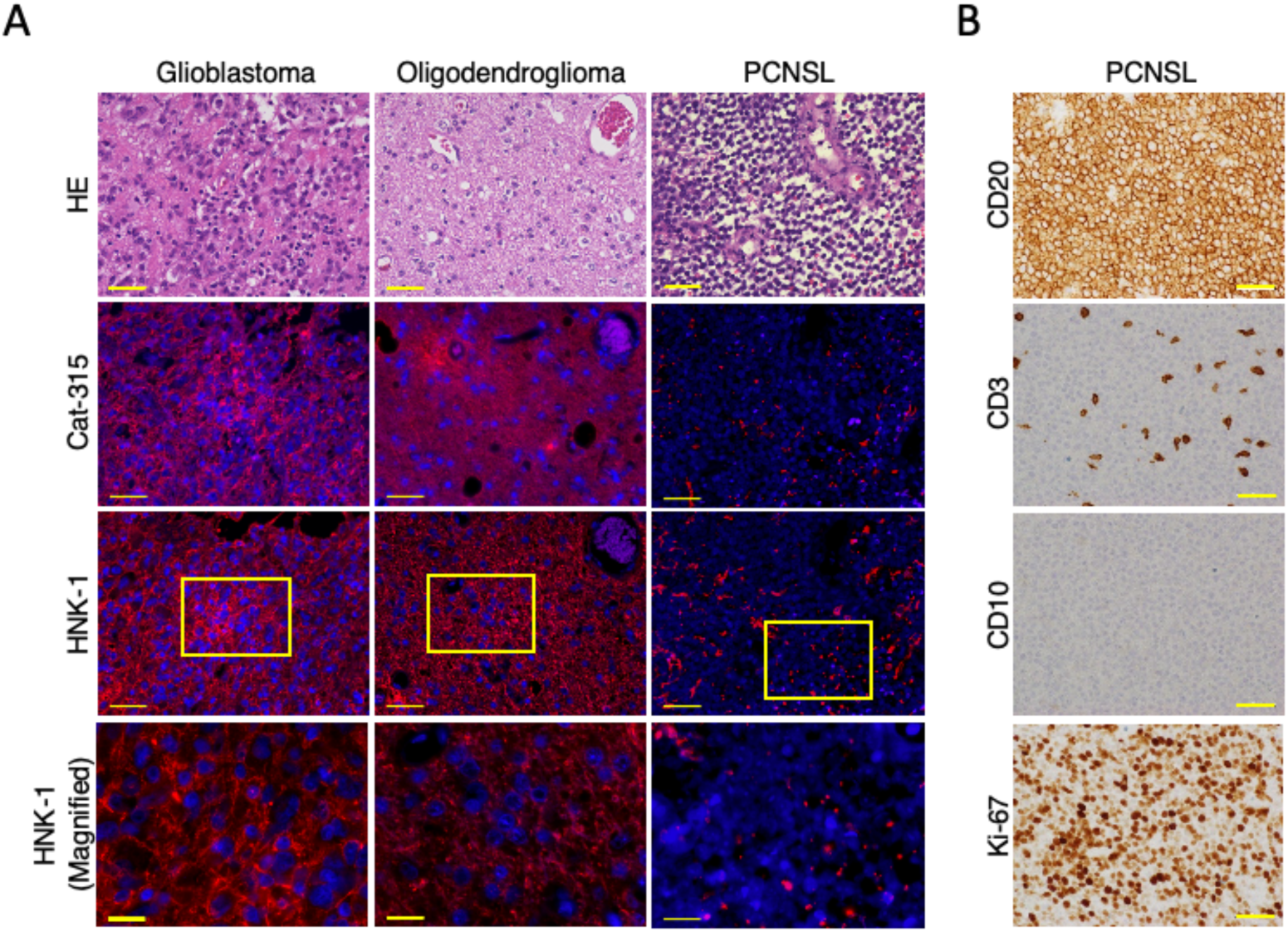
**HNK-1 expression in glioma tissues.** (A) Immunofluorescent images of brain sections from patients with glioblastoma, IDH-wildtype, WHO grade 4, oligodendroglioma IDH-mutant and 1p/19q-codeleted, WHO grade 2, and primary diffuse large B-cell lymphoma (not included in CSF analysis). Sections were stained with the Cat-315 (red) or HNK-1 (red) antibodies, and for DAPI (blue). Scale bar: 50 µm or 20 µm (magnified area). (B) Immunohistochemical images of primary diffuse large B-cell lymphoma stained for CD20, CD3, CD10, and Ki-67. Scale bar: 50 µm.

### Analysis of PTPRZ RNA levels in glioma and PCNSL cells

The detection of sPTPRZ in CSF and immunohistochemical analysis we have conducted both rely on antibodies that recognize the HNK-1 glycan chain of PTPRZ. The absence of PTPRZ in PCNSL cannot rule out the possibility that PTPRZ is expressed but lacks HNK-1 glycan chains. We then examined the mRNA expression levels of PTPRZ and the brain-specific glycosyltransferases GnT-IX, involved in the HNK-1 epitope formation, in glioma and PCNSL (Fig. 3A). Since PCNSL tumor samples are typically collected in minimal amounts solely for diagnostic purposes, sufficient tissue for conducting transcriptome analysis is not available.

**Figure 3.**
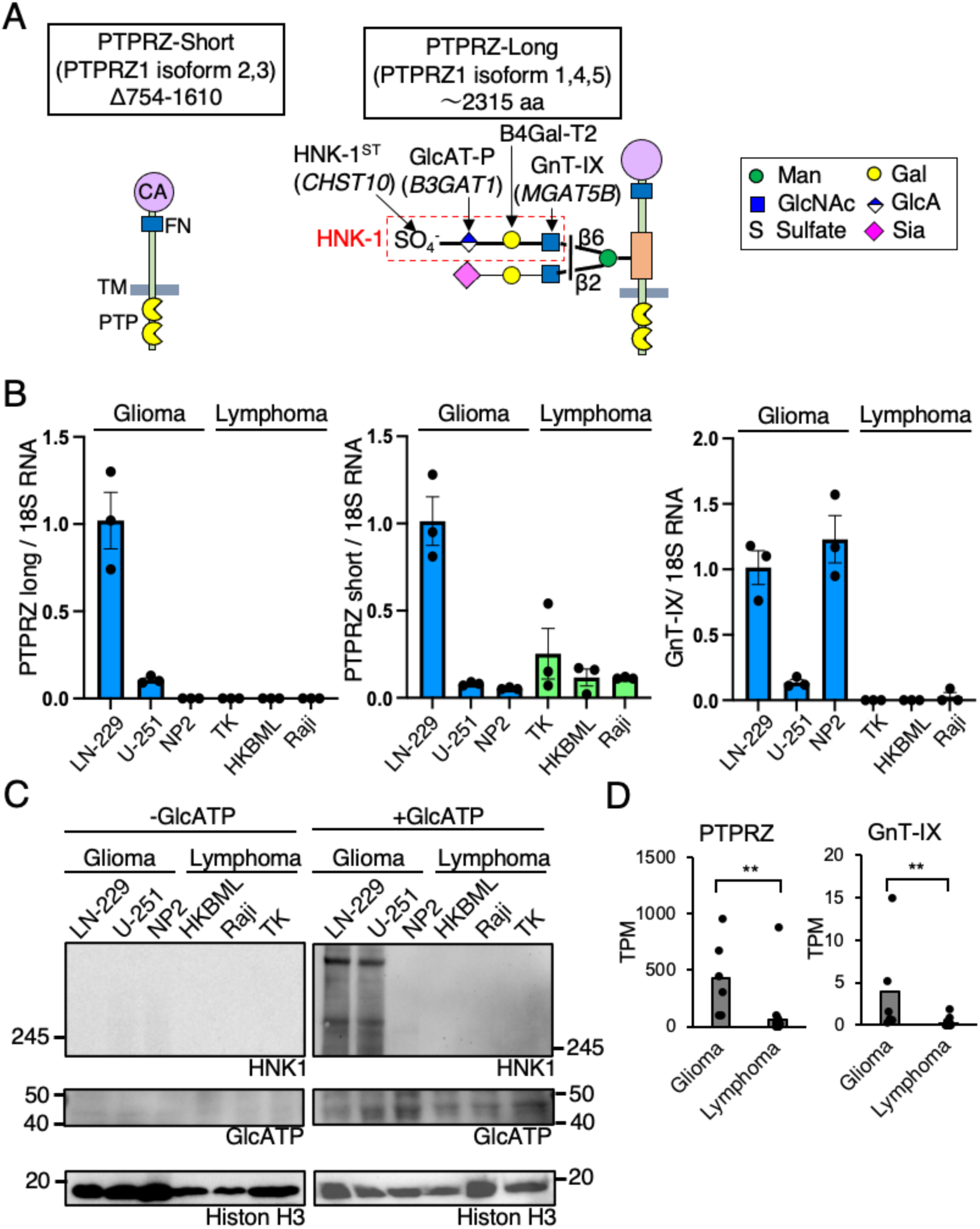
**PTPRZ mRNA and protein expression in glioma and PCNSL.** (A) Schematic representation of PTPRZ isoforms and their alternative splicing patterns. (B) Quantitative PCR analysis of PTPRZ mRNA expression in glioma (LN-229, NP2, U-251) and lymphoma-derived (TK, HKBML, Raji) cell lines. (C) Western blot analysis of PTPRZ protein expression in glioma and lymphoma cell lines with and without GlcAT-P transduction. (D) RNA-seq analysis comparing PTPRZ and GnT-IX mRNA expression in glioma and PCNSL tumor tissues from the GEO database. Data are presented as mean ± SEM. Statistical analysis was performed using the Mann–Whitney U test.

Therefore, we first performed qPCR analysis using cultured cell lines derived from glioma (LN-229, NP2, and U-251), PCNSL (TK, HKBML), and lymphoma (Raji). PTPRZ has multiple mRNA isoforms due to alternative mRNA splicing, and in humans, there are two main groups: PTPRZ-long and PTPRZ-short (Fig. 1A)^26^. High level of CSF sPTPRZ in glioma patients is derived from PTPRZ-long^9^. We found that both LN-229 and U-251 cells expressed significant levels of PTPRZ long form mRNA, whereas it was barely detectable in NP2 cells (Fig. 3B). In the PCNSL and lymphoma-derived cell lines, expression of PTPRZ-long form was undetectable. While, PTPRZ-short mRNA is expressed at higher level in LN-229 and at a lower level in other cell types. GnT-IX eoncoding MGAT5B was expressed in all glioma-derived cell lines examined, but was not detected in PCNSL or lymphoma-derived cell lines. Next, we analyzed the PTPRZ protein levels in these cultured cell lines. It has previously been shown that mRNA expression of GlcAT-P, which is essential for the synthesis of the HNK-1 epitope, is significantly reduced *in vitro* cell culture conditions^16^. Therefore, each cultured cell line was transduced with a retroviral vector to achieve ectopic expression of GlcAT-P, enabling the detection of PTPRZ using the HNK-1 antibody. Indeed, without GlcAT-P expression, PTPRZ was undetectable in the lysates from all cell lines, including glioma (Fig. 3C). When GlcAT-P was expressed, PTPRZ protein was detected in glioma cell lines LN229 and U-251; however, it was not detectable in glioma NP2 cells or in any of the lymphoma cell lines, consistent with the qPCR results. Furthermore, RNA-seq data of tumor tissues derived from glioma (GSE266210) and PCNSL (GSE155398) were obtained from GEO, and we found that both *PTPRZ1* and *MGAT5B* mRNA levels were significantly higher in glioma samples compared to PCNSL samples (Fig. 3D). Taken together, these findings indicate that glioma exhibits markedly higher PTPRZ mRNA expression compared to PCNSL. Additionally, GnT-IX, previously reported as a therapeutic target for glioma^16^, was also found to be highly expressed in glioma.

### Quantitative analysis of sPTPRZ in the CSF among Glioma, PCNSL and Control patient

Several lines of data shown in Figure 1 indicate that the anti-HNK-1 antibody possesses high specificity and sensitivity for detecting CSF sPTPRZ. We then used the anti-HNK-1 antibody as both the capture and detection antibodies and established a sandwich ELISA system. For the standard, recombinant sPTPRZ754 protein modified with HNK-1 glycan was used^16^, yielding linearity and confirming the assay’s performance (Fig. 4A). Using this sandwich ELISA, we quantified CSF sPTPRZ levels in 86 samples, including 51 glioma cases, 15 PCNSL cases, and 20 control cases (Table I), in which the CSF sPTPRZ levels were significantly higher in glioma patients compared to controls (Fig. 4B), being consistent with Figure 1D. CSF sPTPRZ showed a tendency to be elevated regardless of glioma grade (Fig. 4C), and was also elevated across different glioma subtypes including oligodendroglioma, astrocytoma, and GBM (Fig. 4D), suggesting that sPTPRZ may serve as a general marker for glioma. The receiver operating characteristic (ROC) curve for differentiating glioma from controls showed an area under the curve (AUC) of 0.910. A cutoff value of 0.31 μg/mL provided a sensitivity of 82 % and specificity of 95 % (Fig. 4E). As compared with PCNSL patients, the CSF sPTPRZ levels in glioma were significantly higher; ROC curve for distinguishing between the two groups showed an AUC of 0.811, with a sensitivity of 60 % and a specificity of 86 %, indicating moderate diagnostic accuracy (Figure 4F). It should be noted that 14 % of the CSF samples from glioma patients exhibited sPTPRZ levels below the cutoff value, consistent with findings in glioma cell lines such as NP2, which showed minimal PTPRZ expression.

**Figure 4.**
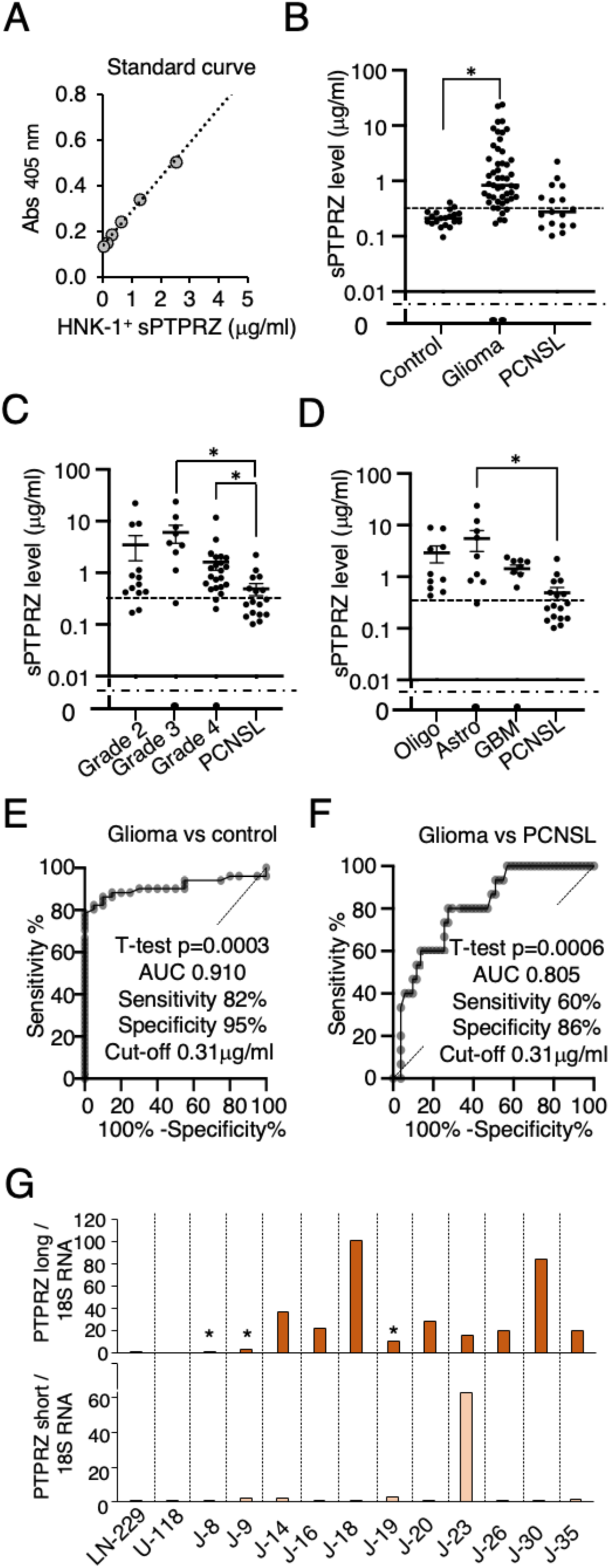
**Quantitative analysis of CSF sPTPRZ in glioma, PCNSL, and control patients using a sandwich ELISA.** (A) Standard curve for the newly developed sPTPRZ ELISA, demonstrating linearity of the assay. (B) CSF sPTPRZ levels in control (n=20), glioma (n=51), and PCNSL (n=15) patients. The horizontal dotted line indicates the ELISA detection limit, and the dashed line represents the cutoff value (0.31 µg/ml). *p < 0.05 (Kruskal–Wallis test with Dunn’s post hoc test). (C) sPTPRZ levels among glioma subtypes (Grade 2, Grade 3, Grade 4 gliomas) and PCNSL. *p < 0.05. (D) Comparison of CSF sPTPRZ levels among oligodendroglioma (Oligo), astrocytoma (Astro), glioblastoma (GBM), and PCNSL. *p < 0.05.(E) ROC curve analysis comparing glioma and control groups. (F) ROC curve analysis comparing glioma and PCNSL groups. (G) Quantitative PCR analysis of long and short isoforms of PTPRZ mRNA normalized to 18S rRNA in glioma tissues. The asterisks indicate samples in Fig. 4B that had sPTPRZ ELISA values below the cutoff threshold. Statistical analysis was performed using the Welch’s t-test. ROC curves in (E) and (F) were generated using Prism 10.5.

We next performed qPCR analysis on glioma samples with high CSF sPTPRZ levels (n=8) and those with levels below the cutoff (n=3) , including LN-229, a cell line known for relatively high PTPRZ expression (Fig. 3B), and U-118, in which a PTPRZ-MET gene fusion has been reported ^27^. In glioma samples with high CSF sPTPRZ levels, the average PTPRZ mRNA expression was 40 times higher compared to LN-229 (Figure 4G). In contrast, glioma samples with CSF sPTPRZ levels below the cutoff exhibited significantly lower PTPRZ expression.

### PTPRZ-MET gene fusion protein lacks HNK-1 epitope region

Recently, accumulating reports have described glioma cases with several types of PTPRZ gene fusion, in which most of PTPRZ protein part is replaced with oncogenic proteins, such a MET, ETV1, and BRAF ^27,28–30^. Although the presence of the HNK-1 epitope depends on its site of addition, gene fusion products that lack most of the PTPRZ mRNA are likely to be PTPRZ-negative. To determine the localization of the HNK-1 epitope on non-fusion sPTPRZ, we performed site mapping using mass spectrometry. sPTPRZ was immunopurified from glioma patient CSF using an anti-HNK-1 antibody, digested with endoproteinase Lys-C followed by trypsin, and the enriched glycopeptides were treated with peptide-N-glycosidase F. O-glycopeptides were captured again and used for LC/MS analysis. Glycopeptides were identified from their higher energy collision-induced dissociation spectra by database search using Byonic with a glycan DB including GlcA-containing glycan compositions. Two glycopeptides, DGSVTSTKLLFPSK (1384-1397, Figure S1) and ATSELSHSAK (1398-1407, Figure 5A) were found to have GlcA-containing glycan, HexNAc(2)Hex(2)GlcA(1)Sulfo(1) and HexNAc(1)Hex(2)GlcA(1)Sulfo(1), respectively. Their spectra indicate that hexose is attached first to each peptide and further extension of HexNAc-Hex-HexA-sulfate occurred at both sites. Presumed glycan structures are shown in the inset of the figures. The result indicates that O-Man glycans capped with HNK-1 epitope attach to exon 12, correspond to the juxtamembrane region unique to the long isoform. This finding aligns with our previous observation that the short isoform of sPTPRZ is not detected by Western blotting using anti-HNK-1 antibody^9^. Next, we investigated PTPRZ gene fusion in glioma samples from the C-CAT database (GenMine TOP cancer genome profiling system). Analysis of 301 glioma samples identified 6 *PTPRZ1* gene fusion cases, all of which occur at the *MET* and *PTPRZ1* loci (Figure 5B, Figure S2), indicating that the occurrence of PTPRZ-MET gene fusion in glioma is relatively high, 2%. All fusion-positive cases were found in primary glioma samples. The

**Figure 5.**
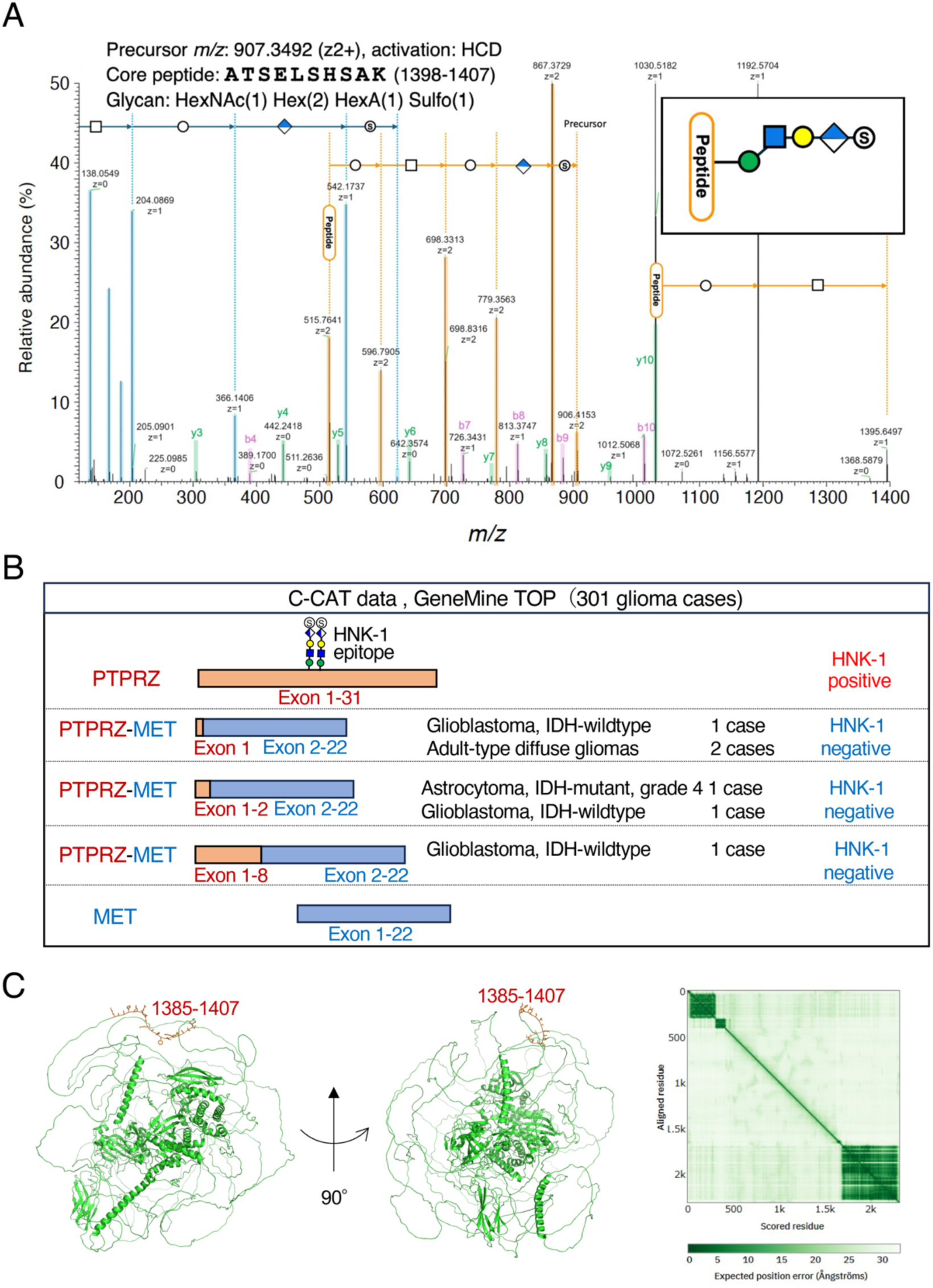
**PTPRZ-MET fusion and HNK-1 glycosylation site mapping.** (A) Annotated tandem mass (MS/MS) spectrum of an HNK-1-carrying glycopeptide, in which core peptide is ATSELSHSAK (1398-1407). After Peptide-N-glycosidase treatment, O-glycopeptides were obtained by HILIC from the digest after and analyzed by LC/MS. (B) Schematic representation of PTPRZ-MET gene fusion variants identified in glioma samples from the C-CAT database. The LC-MS/MS analysis of sPTPRZ shown in (B) revealed that the HNK-1 epitope specifically binds to the juxtamembrane region (exon 12) of the long form of PTPRZ. (C) AlphaFold-based structural model of PTPRZ highlighting the predicted HNK-1 glycosylation sites (1385-1407) in the disordered region.

PTPRZ-MET fusion transcripts were found in both IDH mutated and wild-type gliomas, as has been reported^27^. Multiple types of *PTPRZ-MET* fusion transcripts were observed, with the *PTPRZ1* fusion junction located at exon 1, 2, or 8, while *MET* was consistently fused at exon 2, preserving its kinase domain, as previously described by other groups^27,28^. Since the HNK-1 epitope has only been identified at exon 12 of *PTPRZ1*, none of the fusion products detected in the C-CAT database are expected to carry the HNK-1 epitope and are therefore considered PTPRZ-negative.

## Discussion

Differentiating gliomas from PCNSL and other mimicking conditions such as multiple sclerosis (MS) remains a clinical challenge. Despite advancements in imaging technologies, these diseases often present with overlapping radiological features, and definitive diagnosis still requires invasive tissue biopsy. However, since treatment paradigms for gliomas and PCNSL differ fundamentally, there is a clear need for reliable, minimally invasive biomarkers. In recent years, frequent mutations in MYD88 and CD79B have been identified in PCNSL^31^.

Consequently, liquid biopsy using CSF or blood to check these mutations has garnered increasing attention as a method for differentiating PCNSL^32^, and they will be promising biomarkers for diagnosis of PCNSL. It is important to underscore that the primary aim of this study is the development of a novel biomarker specifically for gliomas. In this context, particular attention is given to evaluating the diagnostic performance of CSF sPTPRZ in distinguishing gliomas from PCNSL.

Previous studies have demonstrated elevated CSF sPTPRZ levels in glioma patients, distinguishing them from controls and MS^9^. However, the detection methods employed in those studies relied on IgM antibodies requiring enzymatic digestion, limiting their clinical applicability. In contrast, the anti-HNK-1 IgG antibody used in our study allows robust detection of sPTPRZ without chondroitinase treatment, enabling the development of a clinically practical ELISA system.

Our data showed that sPTPRZ levels were significantly higher in glioma than in PCNSL or control groups, supporting its role as a glioma-specific biomarker. Notably, the ELISA assay demonstrated high sensitivity (82%) and specificity (95%) for distinguishing gliomas from controls. While its performance in differentiating gliomas from PCNSL was moderate (sensitivity = 59%), specificity remained high (86%), suggesting that CSF sPTPRZ may be particularly useful for pre-biopsy screening or patient triage. When used in combination with other modalities such as MRI, it is expected to enhance diagnostic accuracy by providing corroborative evidence and facilitate timely planning of surgical strategies and patient management.

Importantly, a subset of glioma patients exhibited low CSF sPTPRZ levels, which initially appeared inconsistent with the overall diagnostic trend. Although we could not directly confirm PTPRZ-MET fusions in these particular cases due to an insufficient number of tumor samples available for RNA analysis, analysis of glioma datasets revealed that such fusions are present in a distinct subset of tumors. Several types of PTPRZ-MET gene fusions exist, generating fusion proteins in which most of the MET kinase domain is preserved^27,33^. Our mass spectrometry analysis of purified sPTPRZ from CSF demonstrated that the HNK-1-capped O-mannose glycan is localized to the juxtamembrane region, which is unique to the PTPRZ-long isoform and consistently absent in all reported PTPRZ-MET fusion variants. This molecular detail offers a plausible explanation for sPTPRZ negativity in some gliomas and reinforces the biomarker’s specificity for intact PTPRZ-long.

To further visualize the region of HNK-1 modification, we generated an AlphaFold model of PTPRZ (Figure 5C)^34,35^. The model revealed that the modification sites reside in a highly disordered region of the protein, further supporting the hypothesis that the epitope is unique to the full-length form of PTPRZ.

Interestingly, MET inhibitors have shown therapeutic efficacy in tumors with MET fusions in other cancers, such as lung adenocarcinoma^36^. The presence of PTPRZ-MET fusion in gliomas suggests a similar therapeutic opportunity, as indicated in recent studies^37^. Thus, detecting fusion-positive gliomas through CSF profiling could serve not only as a diagnostic tool but also to guide targeted therapy.

Additionally, our findings confirm that PTPRZ and its glycosylation machinery— especially GnT-IX—are selectively expressed in gliomas but absent in PCNSL. This supports a glioma-specific glycosylation signature, which may contribute to tumor pathophysiology and therapeutic resistance. Targeting such pathways, as shown with GnT-IX inhibition in previous studies^16^, may present novel therapeutic opportunities.

Lastly, our previous data show that CSF obtained via lumbar puncture is suitable for sPTPRZ quantification^9^, broadening the test’s application beyond intraoperative settings. This finding emphasizes the feasibility of using CSF sPTPRZ as a minimally invasive biomarker for glioma diagnosis and disease monitoring in outpatient clinical practice.

In conclusion, CSF sPTPRZ, as detected using an anti-HNK-1 antibody, serves as a highly specific and minimally invasive biomarker for gliomas. It reliably distinguishes gliomas from PCNSL and holds promise for identifying PTPRZ-MET fusion-positive cases. Future prospective studies are warranted to validate these findings and facilitate clinical translation toward more personalized treatment strategies.

## Methods

### Sex as a biological variable

This study involved both male and female patients. Sex was not considered as a biological variable in the study design, and analyses were not stratified by sex. The findings are expected to be relevant to both sexes.

### Ethics

This study was approved by the ethics committee of Fukushima Medical University (approval numbers 2020-287 and 29378), which is guided by local policy, national laws, and the World Medical Association Declaration of Helsinki.

### Human samples

CSF samples and tumor specimens were collected at Fukushima Medical University, Tokyo Women’s Medical University, and Kobe University. CSF samples were collected during surgery without deviating from the normal procedures. After removal of cells and debris, CSF samples were stored at −80℃ until analysis. Tumor specimens collected upon biopsy or tumor removal, fixed with formalin, embedded in paraffin and sliced into 4-µm-thick sections for Hematoxylin and Eosin or immunohistochemical staining. Tumors were diagnosed and graded according to the current WHO Classification^3^. The control subjects were nontumor disorders including idiopathic normal pressure hydrocephalus, unruptured cerebral aneurysms, facial spasm, or trigeminal neuralgia. The clinical profiles of the patients are summarized in Table 1.

**Table 1.** Clinical characteristics and diagnostic classification of study cohort by tumor or disease type.

| **Tumor/disease type** | **No. of patients** |
| --- | --- |
| **Glioma** | **51** |
| Glioblastoma, IDH-wildtype, WHO grade 4 | 16 |
| Oligodendroglioma, IDH-mutant and 1p/19q-codeleted, WHO grade 3 | 2 |
| Oligodendroglioma, IDH-mutant and 1p/19q-codeleted, WHO grade 2 | 10 |
| Astrocytoma, IDH-mutant, WHO grade 4 | 2 |
| Astrocytoma, IDH-mutant, WHO grade 3 | 7 |
| Astrocytoma, IDH-mutant, WHO grade 2 | 2 |
| Diffuse pediatric-type high-grade glioma,<br>H3-wildtype and IDH-wildtype, CNS WHO grade 4 | 2 |
| Diffuse midline glioma, H3 K27-altered, CNS WHO grade 4 | 1 |
| Others | 9 |
| **PCNSL** | **15** |
| Primary diffuse large B-cell lymphoma | 15 |
| **Control** | **20** |
| Unruptured cerebral aneurysm | 7 |
| Trigeminal neuralgia | 5 |
| Hemifacial spasm | 3 |
| Atherosclerotic intracranial arterial stenosis | 3 |
| Idiopathic normal pressure hydrocephalus | 1 |
| Spastic quadriplegic cerebral palsy | 1 |

### Materials

Anti-SGPG (HNK-1) mouse monoclonal IgG antibody (A2706) is a generous gift from TCI Chemicals. The following antibodies were purchased: anti-PTPRζ (catalog no.: sc-33664; Santa Cruz Biotechnology, referred to as “anti-PTPRZ [Santa Cruz]”), Cat-315 (catalog no.: MAB1581;Merck Millipore) mouse IgM, anti-transthyretin (ab9015; Abcam, referred to as “anti-TTR”) polyclonal sheep IgG, anti-3-beta-glucuronosyltransferase 1, (GlcAT-P; catalog no.: ab199156; abcam), anti-CD3 (ready-to-use product (RTU), clone LN10, Leica Biosystems, UK), anti-CD-10 (RTU, clone 56C6, Leica Biosystems, UK), anti-CD20 (RTU, clone L26, Leica Biosystems, UK), anti-Ki-67 (RTU, clone MM1, Leica Biosystems, UK) and anti-histone H3 (catalog no.: H0164; Sigma–Aldrich) rabbit IgG. The following horseradish peroxidase– conjugated secondary antibodies were used: goat anti-mouse IgM (catalog no.: SAB-110; StressGen), and goat anti-mouse IgG (catalog no.: NA931-1ML; Cytiva). The following fluorescently labeled secondary antibodies were used: Alexa Fluor 546 goat anti-mouse IgM (catalog no.:A21045; Life Technologies).

### Cell culture

HEK293FT cells (Thermo Fisher), human glioblastoma LN-229 (CRL-2611; ATCC), U-251 (9063001, Sigma-Aldrich), U-118 (HTB-15; ATCC), anaplastic glioma NP-2 (RCB4498, RIKEN Cell Bank) were maintained in DMEM (D5796; Sigma) containing 10% FBS (Equitech-Bio, Inc). Human primary CNS lymphoma TK (JCRB1206, JCRB Cell Bank), HKBML (RCB0820, RIKEN Cell Bank) and Burkitt’s lymphoma Raji (CCL-86, ATCC) were maintained in RPMI-1640 (R8758; Sigma) containing 10% FBS. Both media were supplemented with penicillin–streptomycin solution (catalog no.: 168-23191; FUJIFILM). GlcAT-P was expressed in a series of glioma and lymphoma cell lines as previously reported^16^.

### Western blot analysis (CSF)

CSF samples (20 µl) were digested with 0.4 mU Chondroitinase ABC (Sigma) in Tris-acetate buffer (pH 7.4) containing protease inhibitor cocktail (Nacalai) for 1 hour at 37℃. The samples were subjected to SDS-PAGE (3-10% gradient gels; Atto) and were transferred to nitrocellulose membranes. After blocking with 5% nonfat dried milk in TBS buffer (pH 7.4) containing 0.1% Tween-20 for 1 hour, the membranes were probed with anti-PTPRZ (Santa Cruz; 1:1000 dilution), anti-HNK-1 (1:1000 dilution), or anti-TTR (1:1000 dilution) antibodies overnight at 4℃, and with the appropriate horseradish peroxidase–conjugated secondary antibodies (1:3000 dilution) for 1 hour at 25℃. The blots were developed using SuperSignal West Femto maximum sensitivity substrate (code: 34096; Thermo Fisher Scientific) for PTPRZ (Santa Cruz) and MET, and ECL Western Blotting Detection Reagent (code: RPN2106; GE Healthcare) for HNK-1 and TTR. Signals were detected with the ChemiDoc Touch MP (Bio-Rad) and quantified using Image Lab Software (Bio-Rad).

### Immunohistochemistry and histology

Paraffin sections of gliomas and PCNSL sliced into 4 µm thickness were obtained from Fukushima Medical University Hospital. The sections were baked at 60℃ for 30 minutes, deparaffinized in xylene, rehydrated in an ethanol series (100%, 90%, 80% and 70%) and heated with Histofine antigen retrieval regent, pH9 (code: 415211; Nichirei biosciences) for 15 minutes, and leaved at room temperature for more than 20 minutes to cool slowly. Following blocked with 5% goat serum in PBS for 15 minutes, sections were incubated with primary antibodies (Cat-315, 1:250 dilution; anti-HNK-1, 1:100 dilution) for 1 hour at 25℃ and with fluorescently labeled secondary antibodies (1:300 dilution) for 45 minutes at 25℃. After washing, VECTASHIELD Vibrance Antifade Mounting Medium (code: H-1800; Vector Laboratories) was applied to sections. Staining for CD20, CD3, CD10 and Ki-67 was performed using standard retrieval on an automated staining machine (Leica BOND-III system, Leica Biosystems, Australia). The Images were captured using a fluorescence microscope BZ-X810 (Keyence).

### Immunoprecipitation (IP)

Digestion of CSF with chondroitinase ABC (Sigma) was performed in phosphate buffer (pH 7.4), as necessary. The CSF samples (20 μl) were incubated with anti-HNK-1 antibody for 2 hour at 4°C on a rotator, and incubated with Dynabeads protein G (DB1003D, Thermo Fisher Scientific) for 30 minutes at 4°C on a rotator. The beads were washed with PBS and used for western blotting. For the purification of CSF sPTPRZ for LC-MS/MS analysis, anti-HNK-1 antibody coupled to Dynabeads protein G with DMP (Thermo Fisher Scientific) was incubated with glioma CSF (2 ml). The sPTPRZ-bound beads were washed with PBS and boiled with 40 μl phase transfer surfactant buffer (PTS-buffer, 12 mM sodium deoxycholate and 12 mM sodium N-lauroylsarcosinate in 50 mM Hepes [pH 8.0])^38^ and the eluate was used for LC-MS/MS analysis.

### sPTPRZ ELISA

An anti-HNK-1 antibody was conjugated with alkaline phosphatase using Alkaline Phosphatase (AP) Conjugation Kit (abcam, ab102850). A 96-well immunoplate (96 well MicroWell MaxiSorp, Nunc) was coated with anti-HNK-1 antibody (10 µg/ml in 0.1M sodium-carbonate buffer, pH9.5) and immobilized overnight at 4℃. After washing the plates with PBS two times, the plate was incubated with 1% BSA in PBS and immobilized overnight at 4℃. After washing the plates with PBS five times, recombinant sPTPRZ754 bearing the HNK-1 epitope was used as a standard^16^, and CSF samples diluted 4-fold with EIA buffer (27736D, IBL) were added.

The plate was then incubated at 37℃ for 30 minutes. After washing the plates with PBS eight times, AP-conjugated anti-HNK-1 antibody (1 μg/ml) was incubated for 30 minutes at 37℃. As a AP colorimetric substrate, 1-Step PNPP (37621; Thermo Fisher Scientific) was used. The optical density was measured at 405 nm (OD_405_) using Nivo 3F plate reader (Perkin Elmer Victor).

### Real time PCR analysis

Glioma tissues were homogenized with 4 ml of TriPure Isolation Reagent (Roche, 11667165001) with gentleMACS Dissosiator (RNA_02 program, Miltenyl Biotec). The RNA pellets were suspended with PBS and further purified with a High Pure RNA Isolation Kit (Roche, 11828665001). For the cells cultured *in vitro*, only a High Pure RNA Isolation Kit was used for RNA isolation. RNA samples (up to 5 μg) were then reverse-transcribed with random hexamers using a Transcriptor First-Strand cDNA Synthesis Kit (Roche, 04379012001) as per the manufacturer’s protocol. The amount of cDNA of specific genes was then quantified using a TaKaRa qPCR probe (TaKaRa) or the Universal ProbeLibrary (Roche) with TaqMan Master (Roche) and a LightCycler 96 system (Roche) in accordance with the manufacturers’ instructions. The primer and probe sequences are shown in Table S2. The levels of mRNA were normalized to the corresponding ribosomal RNA levels and calculated using the comparative cycle threshold (2−Δ Δ Ct) method^39^.

### LC-MS/MS analysis

Enriched sPTPRZ in PTS-buffer was digested with endoproteinase Lys-C (Promega, USA) followed by Trypsin (Promega) after reduction with dithiothreitol (Thermo Fisher Scientific, USA) and carbamidomethylation with iodoacetamide (Thermo). The digests were desalted using GL-Tip SDB (GL Science, Japan) and freeze-dried. The peptides were treated with Peptide-N-glycosidase F (1 mU, Takara, Japan) and O-glycopeptides were captured by hydrophilic interaction chromatography (HILIC) on an Amide-80 column (TOSOH, Japan). Obtained O-glycopeptides were analyzed by LC/MS/MS using UltiMate 3000 RSLCnano–Orbitrap Eclipse tribrid mass spectrometer system (Thermo Fisher Scientific). Peptides were separated using C18 column (inner diameter: 0.075 mm, length: 25 cm, 1.9 μm particle, Nikkyo Technos, Japan) by the gradient of acetonitrile (3-35%, 45 min, 300 nL/min) in 0.1 % formic acid. Eluate was ionized by electrospray method at 2 kV and introduced into the mass spectrometer. Mass spectra were obtained by data-dependent acquisition and positive ion mode.

Both precursor-survey MS and fragment mass (MS/MS) spectra were measured by orbitrap analyzer at the resolution of 120K and 60K, respectively. Selected precursor ions were fragmented by higher-energy collision-induced dissociation method (stepped collision energy of 20, 30, and 40 %). Obtained MS/MS spectra were searched using Byonic (ProteinMetrics, USA) using glycan DB containing 100 glycan compositions including in part HexA or HNK-1 (HexNAc-Hex-HexA-Sulfo). Mass tolerance of MS and MS/MS were 2 ppm and 5 ppm, respectively. Peptide N-terminal pyro-Glutamine (Gln), Peptide N-terminal Ammonia loss (Cys), deamidation (Asn), and oxidation (M) were considered as variable modifications. Protein database used was SwissProt_Human fasta file (downloaded on May 19, 2020, 40,000 entries). The results were processed with Proteome Discoverer (ver.2.5, Thermo Fisher Scientific). The data showing “Confidence”=“High” and glycosylation were treated as “an identified glycopeptide”.

### C-CAT data analysis

We accessed C-CAT database on 29th and 30th January 2025 (in total 88,688 cases were registered on 30th January) and 1938 cases of gliomas were registered in the database. To efficiently collect the PTPRZ1 fusion cases, we selected the cases that were analyzed with GenMineTOP, a comprehensive genomic panel whose RNA-seq data were confirmed superiority on the DNA panel in the detection of fusion genes^40^. We obtained 301 glioma cases and 6 cases showed the PTPRZ1-MET fusion gene products.

### Bioinformatic analysis

Comprehensive gene expression profiles of lymphoma (GEO accession GSE155398^41^) and glioma (GEO accession GSE266210^42^) were obtained from RNA-seq data published in the GEO database^43, 44^. For the quantification of gene expression profiles, we used the NCBI-generated data on which the estimated gene expression abundances were pre-calculated using the pipeline built by the NCBI SRA and GEO teams. Briefly, the RNA-seq data are aligned to the reference genome (GRCh38; GCA_000001405.15) using HISAT2^40^, and the raw counts of those where more than 50% of the reads are aligned are quantified using Subread featureCounts^45^ with *Homo sapiens* annotation data (release 109). The gene expression profile in each sample was quantified as transcripts per million (TPM) from the raw count data. Among the genes whose expression was precomputed, the estimated expressions abundances for PTPRZ and GnT-IX were extracted.

### Statistical analysis

Data are presented as the mean ± SEM. All groups were tested for normality using the Shapiro– Wilk test, and outliers were detected with the Smirnov–Grubbs test. Comparisons between two groups were performed using the Student’s t-test or Welch’s t-test. Multiple comparisons were performed by one-way analysis of variance with the Tukey–Kramer test. All analyses were performed using GraphPad Prism 9.1.2. (Statcon).

## Data availability

The LC-MS/MS data generated in this study have been deposited in jPOST database under accession number [<u>JPST003723</u>/PXD062244]. Other datasets generated or analyzed during the current study are available from the corresponding author S. Kitazume upon reasonable request. Supplementary Material is available at *JCI* online.

## Author contributions

M.F. and S. Kitazume designed the project; Y.N., K.N., R.H., K.H., K. Kanai, S. Koriyama., T. Saito, Y.M., T.K. and T. Sasayama prepared the human samples; Y.N., T. Sakagami., T.K., K.T. J.I., and H.S. performed the biochemical and cellular experiments; Y.N., Y.O., Y.H. and M.U. conducted an immunohistochemical analysis; S.G., H.A. and H.K. performed the LC-MS/MS analysis; J.Y. conducted C-CAT data analysis, K. Kawata conducted a bioinformatic analysis; Y.Y. performed structural analysis; Y. Kawaguchi oversaw the cellular experiment; and Y.N., M.F. and S. Kitazume wrote the manuscript. All authors commented on the manuscript. All authors declare no competing interests.

## Data Availability

The LC-MS/MS data generated in this study have been deposited in jPOST database under accession number [JPST003723/PXD062244]. Other datasets generated or analyzed during the current study are available from the corresponding author S. Kitazume upon reasonable request.

## Acknowledgments

This work was supported by AMED (grant numbers JP22cm0106484 and JP24ama221440), the Mizutani Foundation for Glycoscience (to S.K.), and a grant from the International Joint Research Project of the Institute of Medical Science, the University of Tokyo, and by the joint research program of the J-GlycoNet cooperative network (25C012), which is accredited by the Minister of Education, Culture, Sports, Science and Technology, MEXT, Japan as a Joint Usage/Research Center.

