## Supporting Information for "HNK-1 Antibody-Based ELISA for Soluble PTPRZ: A Practical Cerebrospinal Fluid Biomarker for Glioma"

**Supplementary Table 1.**

**Sequences of qPCR primers and probes used in this study**

| **Targeting gene (Protein)** | **Primer and probe sequence (5'-3')** |
|---|---|
| *PTPRZ1* (PTPRZlong) | F: GACTCAGAAATAACTCCTGGATTCC<br>R: GACCAATACGAGACTCATGGCTA<br>FAM-CCTCTGCCTCTGAAACGTGGAACACTTCTG-BHQ-1 |
| *PTPRZ1* (PTPRZshort) | F: TCCTCCAGACAACAGGATTTGG<br>R: TGGCTACTATTACTGGCCTCATTG<br>FAM- ACGGTCAACGTGGTATACTCGCAGACAACC-BHQ-1 |
| *MGAT5B* (GnT-IX) | F: ACCCTACGAGTACACCTGCG<br>R: GGTAGGGCAGGGTCTGGAG<br>FAM-CACGCCTACATCCAGCACCAGGACTTCT-BHQ-1 |
| *18S ribosomal RNA* | F: GCAATTATTCCCCATGAACG |
| | R: GGGACTTAATCAACGCAAGC |
| | ProbeLibrary probe 48 (Roche) |

The probes for PTPRZ-long (PTPRZ1 isoforms 1, 4, and 5), PTPRZ-short (PTPRZ1 isoforms 2 and 3) and *MGAT5B* genes were labeled with the fluorescent reporter dye FAM at its 5′ end and the quencher dye BHQ-1 at its 3′ end. The probe for ribosomal RNA was labeled with VIC at its 5′ end and the quencher dye BHQ-1 at its 3′ end.

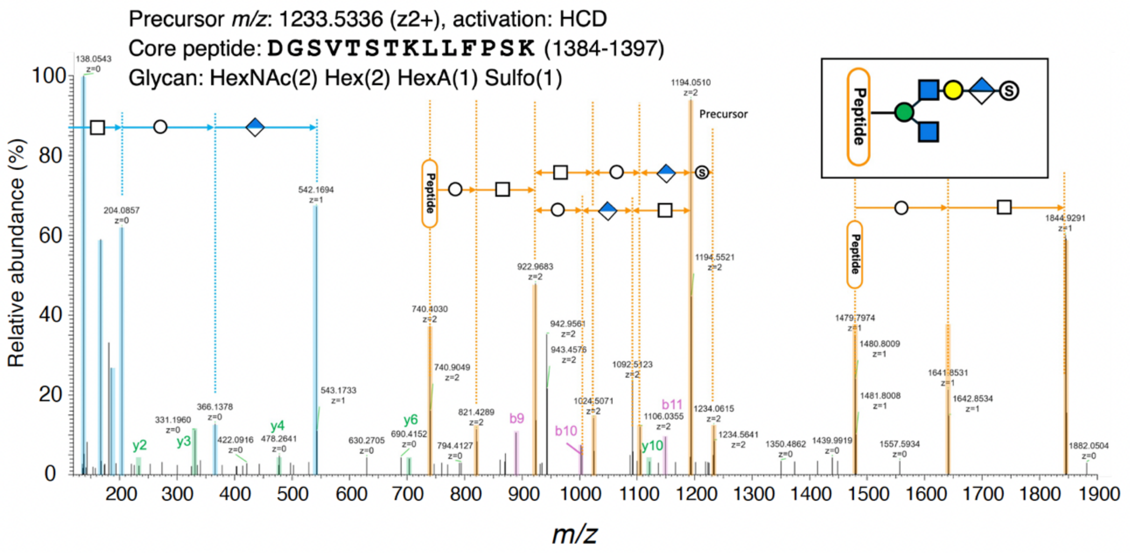


**Figure S1. Annotated tandem mass (MS/MS) spectrum of an HNK-1-carrying glycopeptide.**

The core peptide is DGSVTSTKLLFPSK (residues 1384–1397). Y-ions and B-ions (oxonium ions) generated by glycosidic bond dissociation are annotated and highlighted in blue and orange, respectively. Due to the presence of multiple potential O-glycosylation sites (Ser/Thr) within the peptide and extensive fragmentation of the glycan portion from the peptide backbone, the exact glycosylation site could not be determined. The peptide backbone fragments are shown as b-ions (purple) and y-ions (green).

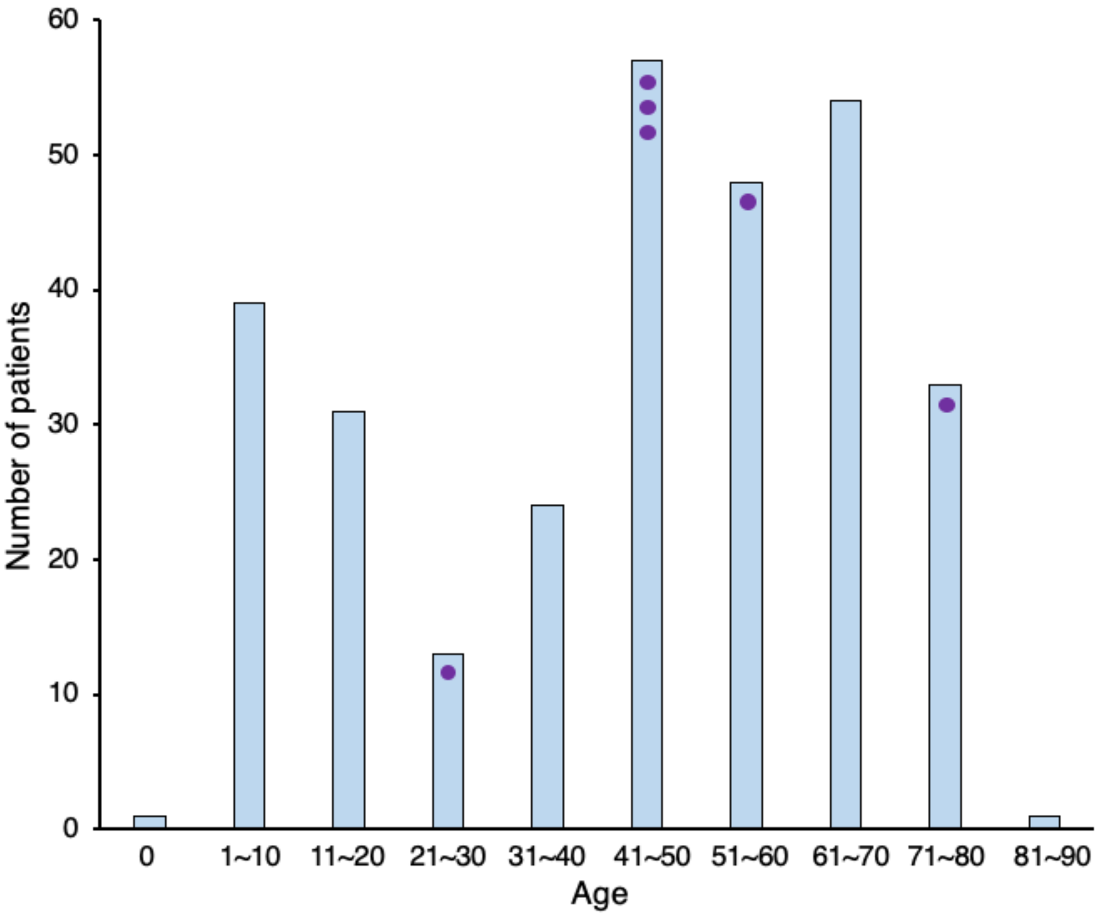


**Figure S2. Age distribution of glioma patients from C-CAT data (GeneMineTOP).**

Blue bars represent the number of patients in each age group. Purple dots indicate individual cases in which the *PTPRZ1-MET* gene fusion was detected.
